# CORRELATION BETWEEN EARLY ANTIBIOTIC THERAPY AND IN-HOSPITAL MORTALITY IN PATIENTS WITH COMMUNITY ACQUIRED INFECTION

**DOI:** 10.1101/2020.05.29.20116848

**Authors:** Beatriz Araújo, Geovane Rossone Reis, Isadora Araújo, Marcella Soares Carreiro Sales, Janne Marques Silveira

**Author notes:** corresponding author: Beatriz Araújo.

## Abstract

Sepsis is defined as the systemic response to an infectious disease, whether caused by bacteria, viruses, fungi or protozoa. Sepsis has several etiologies according to the worsening of the disease, those of bacterial origin with 70%, of which Gram-negative bacilli are the most frequent, especially in patients with the most severe disease. The most common community infections are respiratory infections, urinary tract infections and skin infections, with elderly individuals and children being more susceptible to complications of the immune system. The objective of this research is to analyze whether the time between hospitalization and the start of antibiotic treatment is directly related to the mortality rate and length of stay. This study was carried out in the form of documentary field research, where data collection was performed using data from the medical records of patients diagnosed with community infection or sepsis. Data collection was carried out from a form where patient identification, age, infectious focus, date and time of admission to the emergency room, date and time of the 1st dose of antibiotic, time interval between admission and antibiotic administration, ICU stay, days of mechanical ventilation and outcome. 34 patients were followed up in the emergency room and in the ICU, with an average age of these patients is 71.4 ± 30 years. This research showed a high mortality rate of the included patients who were diagnosed with community infection.

## INTRODUCTION

Sepsis is defined as the systemic response to an infectious disease, whether caused by bacteria, viruses, fungi or protozoa. Manifesting in different clinical stages of the same pathophysiological process^1^. Sepsis symptoms are easy to be confused with other diseases, which makes early diagnosis very difficult^2^. The most common foci where infections occur in the body are: lungs (pneumonia), abdomen (post-surgical periods, biliary and liver infections), kidneys and bladder (urinary and renal infections), skin (wounds, cellulite, erysipelas, openings for the introduction of catheters and probes, abscesses) and in the central nervous system (meningitis)^3^.

According to Siqueira-Batista et al.^4^, sepsis encompasses situations in which systemic inflammatory response syndrome (SIRS) is established, manifesting itself with suspicion or confirmed infection. From a clinical point of view, the presentation of sepsis is related to the multiple possibilities of interaction between man and microorganisms, distinguishing situations such as infection, SIRS, sepsis, severe sepsis, septic shock and dysfunction of multiple organs and systems.

According to Westphal et al.^5^, severe sepsis and septic shock are part of the main causes of death in patients hospitalized in ICUs (Intensive Care Units). Some epidemiological studies have found that this amount is even higher in Brazil, it was noted that mortality rates on the 28th day of hospitalization averaged 57.1% of patients. An assessment carried out by the Sepsis Survival Campaign (CSS) confirmed a clear reduction in the world average of hospital mortality associated with sepsis (37% versus 30.8%). In Brazil, however, this rate is still 55.6% among hospitals participating in CSS.

A prospective, multicenter cohort study was carried out in 65 hospitals in all regions of Brazil. The study was coordinated by the AMIB Brazilian Fund for Education and Research (AMIB Fund – Brazilian Association of Intensive Medicine) and by the Postgraduate Program at the Federal University of Rio de Janeiro (Medical Clinic – Intensive Care area). The participating hospitals received a notebook to fill in the data and with the relevant instructions. The inclusion period was 30 days, and each patient was followed for 28 days or until his discharge from the ICU or death. All patients hospitalized with sepsis or who developed sepsis during the study period were included. Patients under 18 years of age, rehospitalization in the same period of the study, loss of follow-up due to transfer and ICU stay for less than 24 hours were excluded. Of 140 invited ICUs, 81 (57.8%) participated in the study. The study showed a high mortality from sepsis in the ICUs in our country. Mortality in septic shock is one of the highest in the world. Our patients are more severe and have a longer hospital stay^6^.

In a multicentre observational cohort study, they performed in five private and public ICUs, mixed, from two different regions of Brazil. We prospectively followed 1383 adult patients admitted consecutively to these ICUs from May 2001 to January 2002, until their discharge, 28 days of hospitalization or death. For all patients, the following data were collected on admission to the ICU: age, sex, hospital diagnosis and ICU admission, APACHE II score and associated underlying diseases. During the following days, they looked for SIRS criteria, sepsis, severe sepsis and septic shock, in addition to recording the organ failure sequential assessment score. The infection was diagnosed according to the CDC criteria for hospital infection and, for infection acquired in the community, clinical, radiological and microbiological parameters were used. The mortality rate of patients with SIRS, sepsis, severe sepsis and septic shock increased progressively from 24.3% to 34.7%, 47.3% and 52.2%, respectively. For patients with SIRS without infection, the mortality rate was 11.3%. The main source of infection was the lung / respiratory tract^7^.

According to Monteiro et al.^8^, sepsis has several etiologies according to the worsening of the disease, but the most prominent are those of bacterial origin with 70%, of which Gram-negative bacilli are the most frequent, especially in patients with the most common disease. aggravated. Some situations can compromise the host’s immune response and increase the vulnerability to infections, such as: population aging, invasive procedures, immunosuppressed patients and with the human immunodeficiency virus (HIV), use of immunosuppressive and cytotoxic drugs, malnutrition, alcoholism, diabetes mellitus, transplant procedures, nosocomial and community infections and infections by multidrug-resistant microorganisms to antibiotics.

According to Oliveira et al.^9^ during the period in which the patient is hospitalized, this group of patients is subject to mobilization restrictions, which generates severe damages in the functional results. They usually have a prolonged stay in the ICU and remain on mechanical ventilation. As a consequence, the development of musculoskeletal and neuromuscular complications in about 70% to 90% of patients. Immobilization associated with hospitalization, sepsis and inflammation, catalyze physiological changes that propagate low conditioning.

The adverse effect of prolonged deconditioning of rest in humans, with its associated physical inactivity, was known by Hippocrates, who reported loss of strength and exercise performance after prolonged rest and inactivity. The assumption of the horizontal position of the body offers an opportunity to examine the impact of the reduced hydrostatic pressure gradient on the cardiovascular system, unloading force on muscles and bones and less energy use on the physiological reserve capacity.

The premise that deconditioning of bed rest can be partially explained, regardless of the disease, highlights the importance of early intermittent walking and physical activity that can significantly limit many debilitating effects during hospitalization^10^.

Sepsis means: putrefaction, organic decomposition caused by bacteria and fungi. The body’s response to a varied aggression (trauma, pancreatitis, major burn, systemic infection) with the presence of at least two of the following criteria, Hyperthermia: (body temperature> 37°C) or Hypothermia: (body temperature <35°C), Tachycardia :(HR> 90 bpm), Tachypnea: (RR:> 20 bpm PaCO2 <32 mmHg) and Leukocytosis or Leukopenia: (leukocytosis> 12,000 cells / mm3 or leukopenia <4,000 cells / mm3)^11^. According to Valeiro et al.^12^, the therapeutic strategies adopted, which include tissue reperfusion and control of the infectious focus, prove a result of reduced mortality. The lack of identification of the septic condition prevents the initiation of adequate treatment, resulting in progression to multiple organic dysfunctions, which ends up seriously compromising the prognosis of patients. Afterwards, the search for the detection of SIRS during the routine verification of vital signs can imply the probable diagnosis of a septic condition. Antibiotics are natural or synthetic substances, capable of causing death or inhibiting the growth of fungi and/or bacteria. The development of the first synthetic substance with these properties dates back to 1910, through studies by Paul Ehrlich^13^. The progress of drug therapy has been remarkable since the appearance of the first anti-infectives in the 1930s and 1940s, with pharmacological therapy strongly influenced the reduction of morbidity and mortality throughout the 20th century.

In this period, too, the medication is no longer just an instrument of therapeutic intervention, but has become a complex element – technical and symbolic – in Western society^14^. Antimicrobials are one of the main drugs used in the ICU, but their indiscriminate and prolonged use is one of the main factors involved in the emergence of multi-resistant bacteria, with an increased incidence on all continents^15^.

According to the Brazilian National Health Surveillance Agency (ANVISA), a community infection is one that is diagnosed or is incubating when the patient enters the hospital, as long as it is not related to a previous hospitalization in the same hospital. Differentiating from hospital infection, which is acquired after the patient’s hospitalization, manifesting itself during hospitalization or after discharge, when it is possible to relate it to hospitalization or hospital procedures. The most frequent community infections are respiratory infections, urinary tract infections and skin infections, and elderly individuals and children are more susceptible to complications of the immune system^16^.

The average cost per hospitalization or the daily per hospitalization varied between 2008 and 2017. During the years 2008 and 2012 there were some increases, then in the years 2013 and 2014 there was a significant decrease, stabilizing in the years 2015 and 2016. In 2017, the average cost of hospitalization was R$ 2,900^17^.

Available national data point to high mortality, especially in public hospitals linked to the Unified Health System. A single-day prevalence study in approximately 230 Brazilian ICUs, randomly selected in order to adequately represent the group of ICUs studied, points out that 30% of the ICU beds in Brazil are occupied by patients with severe sepsis or septic shock. This study was conducted by the Latin American Sepsis Institute (ILAS), and the results were published on the ILAS website, which are alarming, with a mortality rate exceeding 50%^1^.

This mortality rate in Brazil increased from 2008 to 2016 when looking at the total number of all regions, in 2008 it was 38.85% and in 2016 46.14%. In 2017, there was a reduction from 46.14% to 45.64%. When analyzing by regions, the ones with the highest number of mortality in the year 2017 went to the northeast region with 46.12% and the southeast region with 49.32%. At the state level, sepsis leads to a high mortality rate in Intensive Care Units (ICUs) in the state of Tocantins. In the last 10 years, the number of hospitalizations has decreased, from 362 in 2008 to 146 in 2017, but with an increase in the mortality rate from 37.02% to 61.64%^17^.

Based on these data and in order to assess possible causes and consequences in the lethality of community infectious diseases, the objective of this research is to analyze whether the time interval between hospitalization and the beginning of antibiotic treatment is directly related to mortality and mortality rates. the length of stay in this group of patients.

## MATERIALS AND METHODS

This study was carried out in the form of documentary field research, where data collection was performed using data from the medical records of patients diagnosed with community infection. The type of population recruited for the study were patients with community infection with or without sepsis, over 18 years old, of both sexes, admitted to the ER (emergency room) or to the ICU of the Regional Hospital of Gurupi-TO. The study exclusion group consisted of patients that the hospital did not authorize the use of medical records or that they did not have complete data.

Data collection was carried out based on patient identification, age, infectious focus, date and time of admission to the ER, date and time of the first dose of antibiotic, time interval between admission and administration of antibiotic, stay in ICU, mechanical ventilation days, outcome and total cost. These data were collected between October 24, 2019 and January 24, 2020, with a daily visit to the hospital, between 6 pm and 7 pm, accounting for a total of 37 patients, where, following the sample criteria, three were excluded.

About the statistical analysis of the correlation data between the collected indicators, they were analyzed by Pearson’s coefficient in order to assess the degree of linear association. After validation, they were compared using the Student’s t test paired using the SPSS® software with a 5% significance level, in order to allow the analysis of the probability of death or discharge to occur at random.

According to Resolution No. 466/12 and Resolution No. 510/16, both from the Brazilian Ministry of Health, this research was only carried out after being approved by the Research Ethics Committee.

## RESULTS

34 patients were followed up in the ER and in the ICU, all diagnosed with community infection on admission. Of these 34 patients, 58.8% refer to males and 41.2% refer to females, the average age of these patients is 71.4 ± 30 years. Since the frequency of greatest infection is among patients aged 71 to 80 years old, totaling 37% of the cases presented, as shown in Graph 1.

**Graph1.**
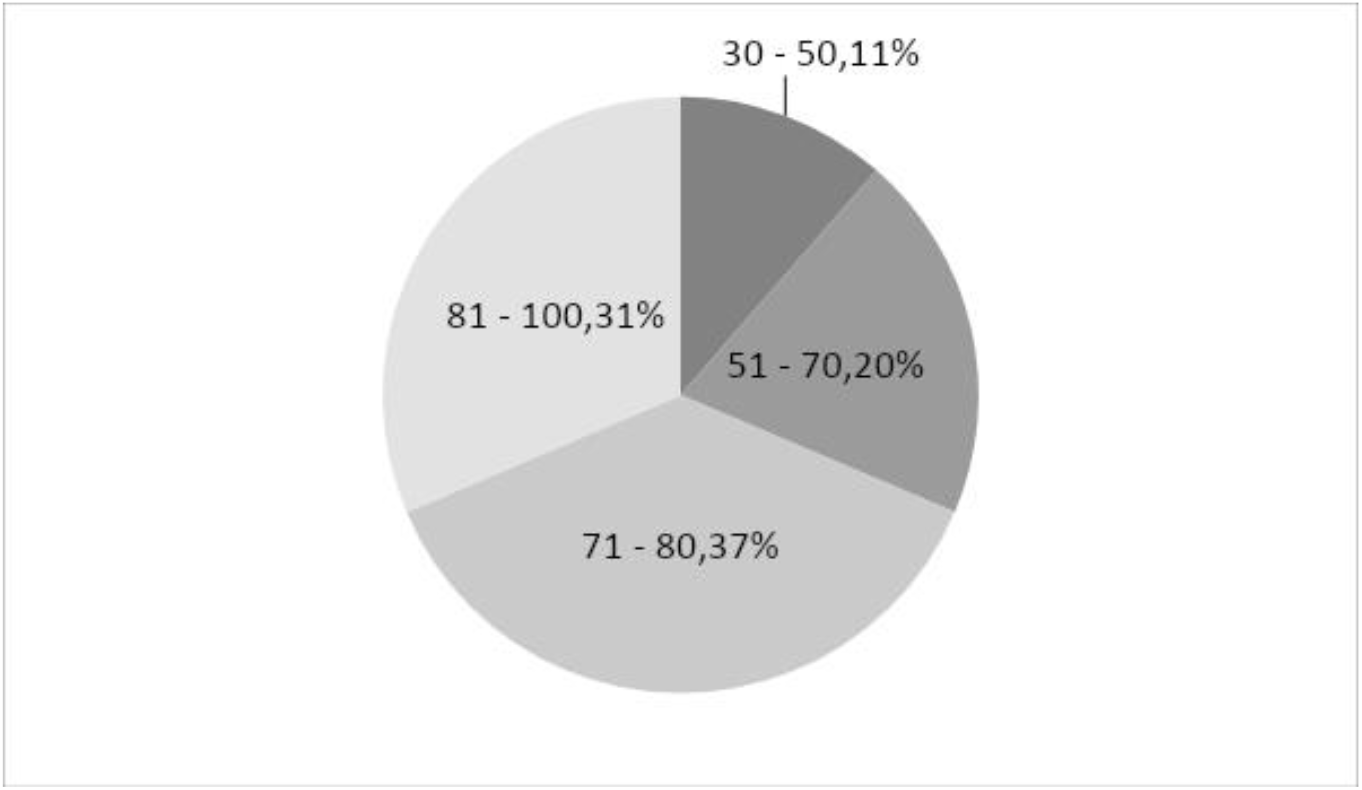
Frequency of greater infection by age ER/ICU, Gurupi-TO, 2019–2020.

In reference to the infectious foci diagnosed during the research period, we found: Furnier syndrome, urinary tract infection, sepsis / septic shock, abdominal infection and pneumonia. The highest prevalence of community infection in the studied period was pneumonia with 76% of the cases admitted to the hospital, as shown in Graph 2. As for the mortality rates due to the corresponding infections cited with 79% of deaths, pneumonia was the most lethal among the infections diagnosed in that period. according to Graph 3. Of these patients 74% were admitted to the ICU and received intensive treatment, with a mortality rate of 72%. Among the patients who were not referred to the ICU and remained in the ER, the mortality rate was 66, 7%, thus totaling 70.6% of death among the patients included in this research.

**Graph 2.**
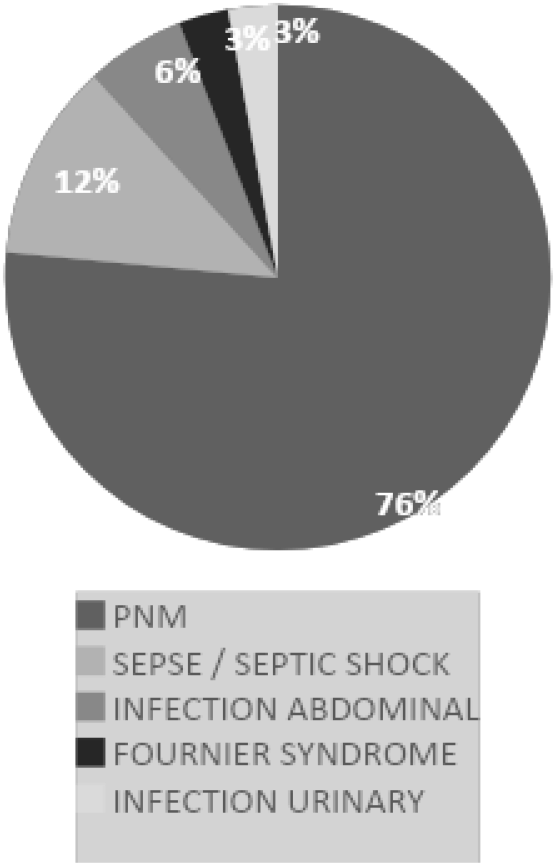
Patients affected with the ER/ICU infectious focus, Gurupi-TO, 2019–2020.

**Graph 3.**
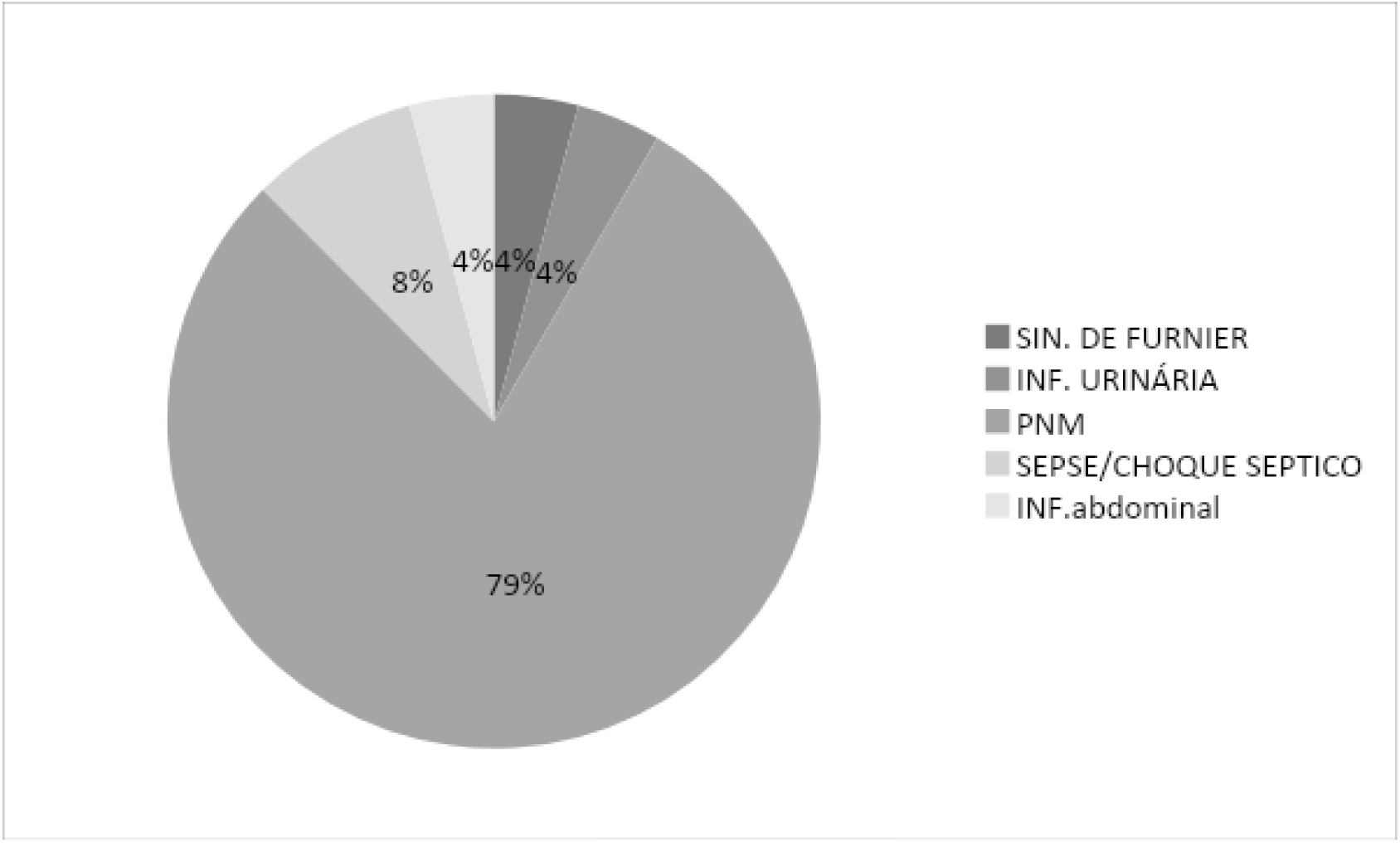
Mortality in infections admitted to the ER/ICU, Gurupi-TO, 2019–2020.

Regarding the use of mechanical ventilation (MV), 91.2% of the patients were intubated, with a mean stay of 14 ± 1 days, of which 77.4% died. The highest mortality rate was concentrated during the first fifteen days of hospitalization of patients with 63% (graph 4). Regarding deaths by length of stay in the PSA and in the ICU, it was observed that both in the ICU and in the PSA the highest mortality rate occurred in the first fifteen days, with an average stay in the ICU of 12.2 ± 1 day.

**Graph 4.**
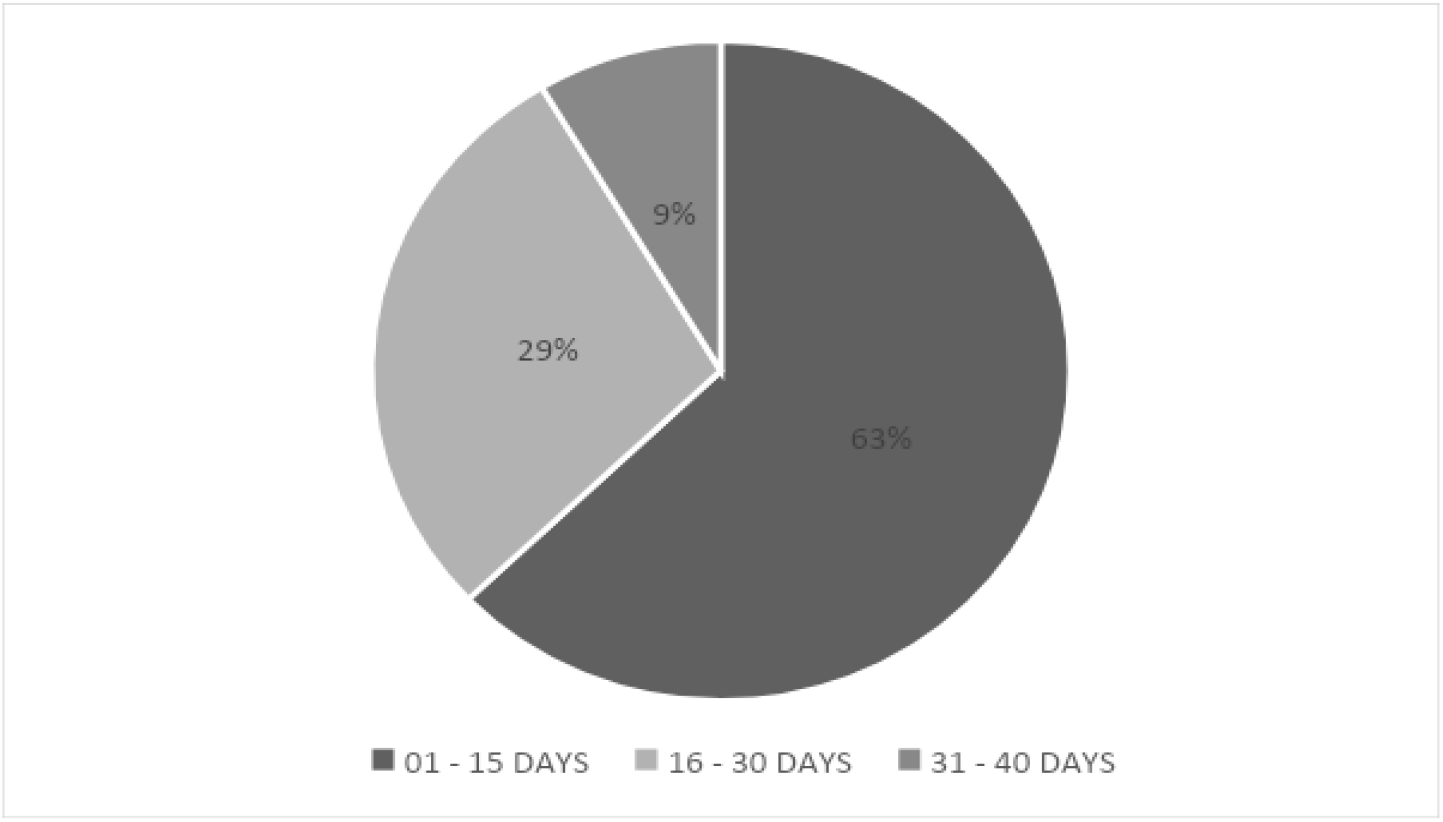
Mortality on days of admission to the ER/ICU, Gurupi-TO, 2019–2020.

Regarding the time interval between admission and the first dose of antibiotic therapy, 26% of patients received it in the first 120 minutes, of which (graph 5) 50% had death as an outcome.

**Graph 5.**
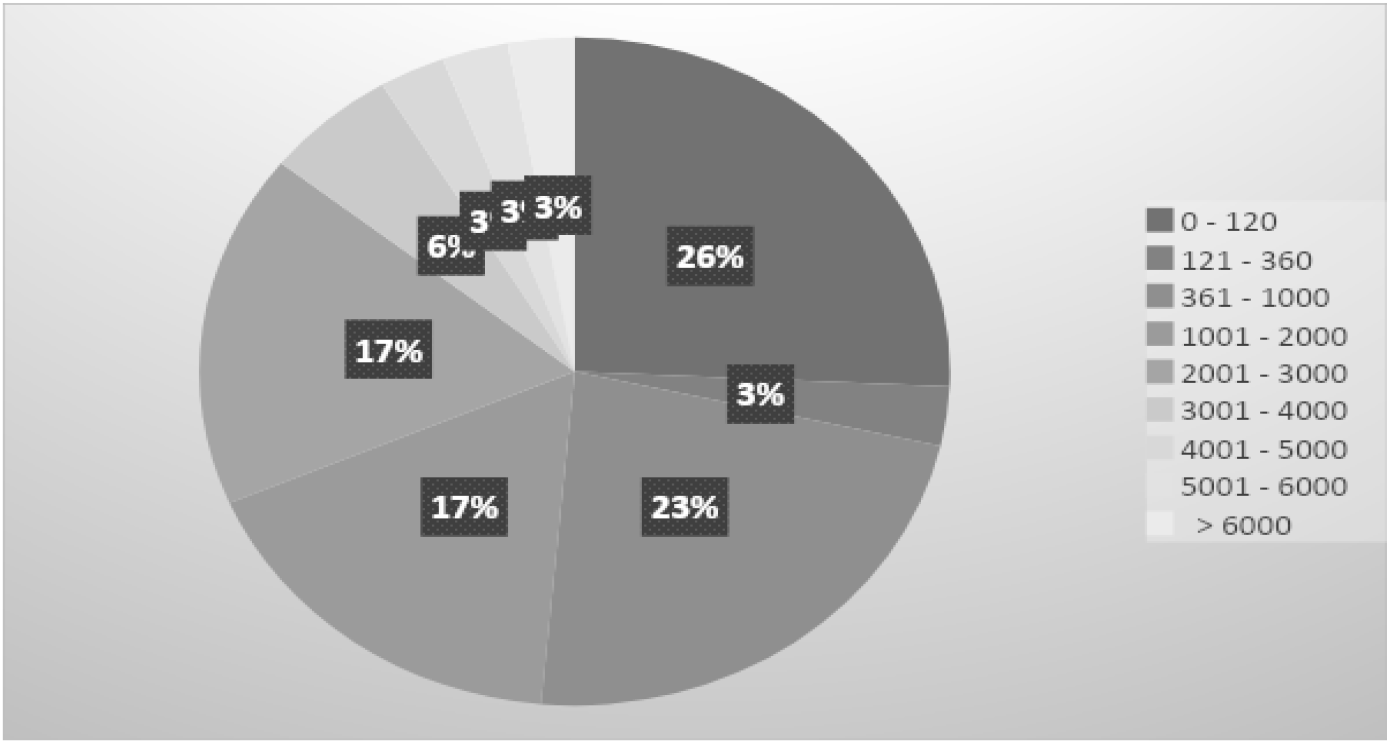
Distribution of patients according to the time interval (in minutes) of admission with the first dose of antibiotic therapy, ER/ICU, Gurupi-TO, 2019–2020.

**Graph 6.**
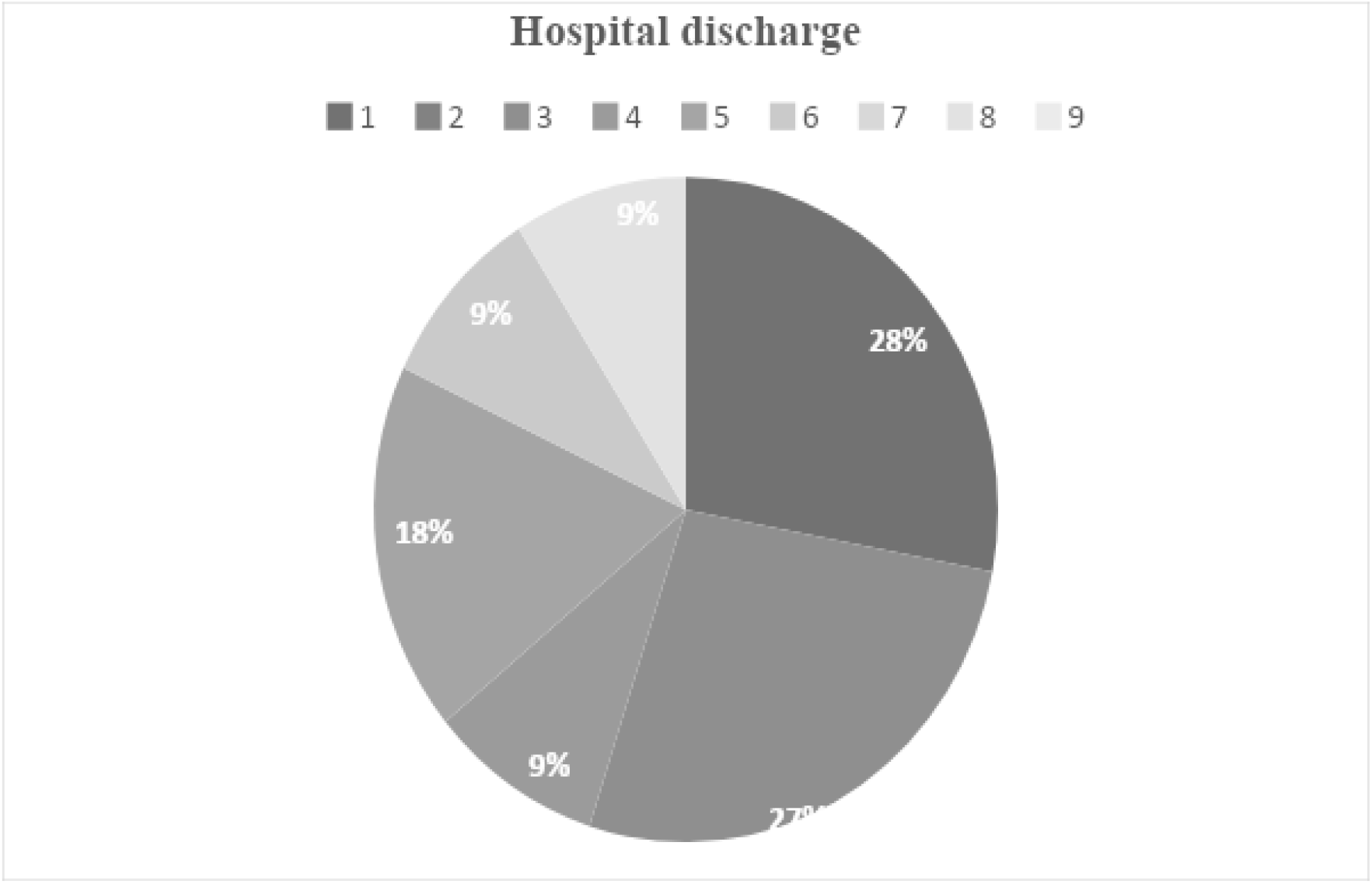
Discharges in hospital stay with beginning of antibiotic therapy, ER/ICU, Gurupi-TO, 2019–2020.

**Graph 7.**
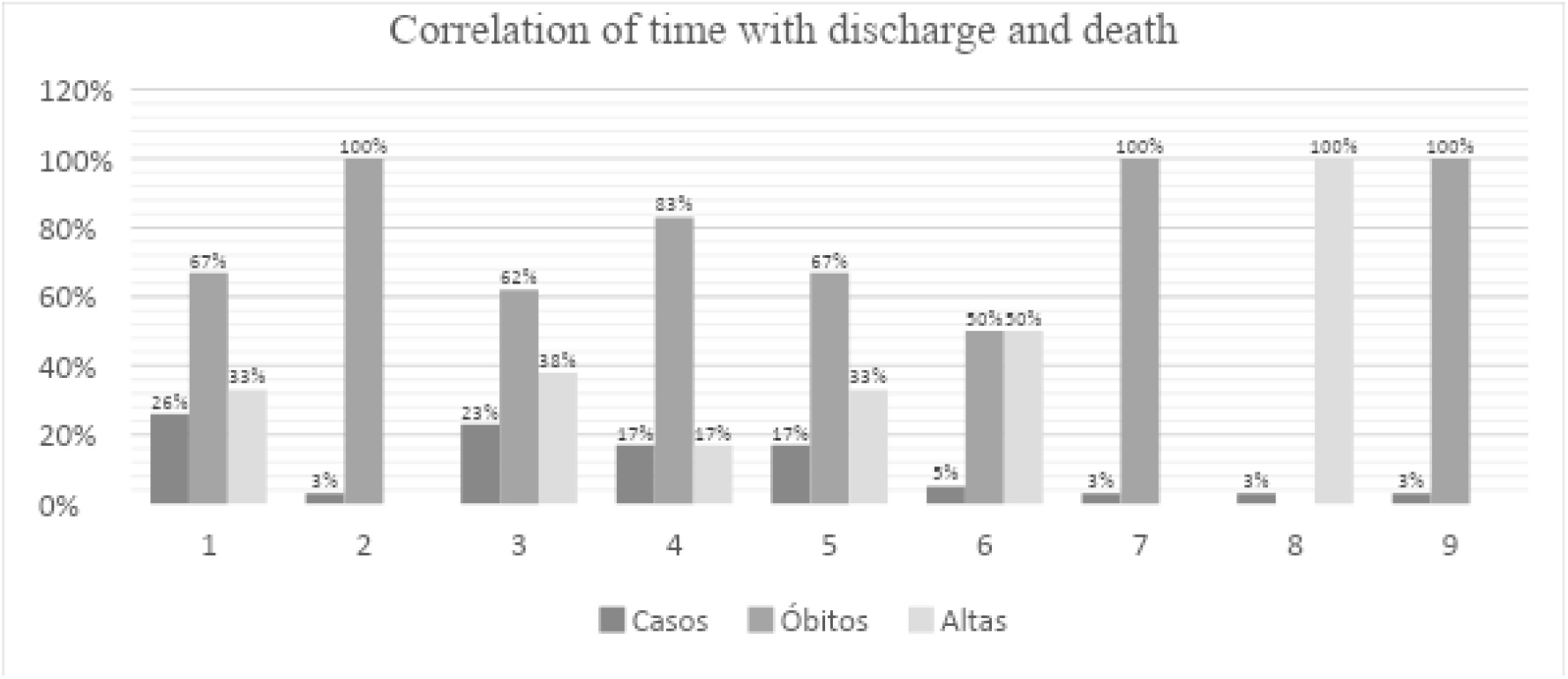
Correlations of antibiotic time with discharge and death, ER/ICU, Gurupi-TO, 2019–2020.

The mortality rate was high in patients who received the first dose of the antibiotic in the first 120 minutes, with 25% of deaths obtained in the study. As shown in Table 1, a quantitative was performed to analyze the time – in minutes – when the largest number of patients hospitalized and included in this study received antibiotic therapy early, in the shortest possible time analyzed, between zero and 120 minutes, as shown in Table 1.

**Table 1.** Quantitative of cases with statistics of death and discharge by time between admission and 1st dose of antibiotic therapy, ER/ICU, Gurupi-TO, 2019–2020.

| **TIME** | **0 - 120** | **121 - 360** | **361 - 1000** | **1001 - 2000** | **2001 - 3000** | **3001 - 4000** | **4001 - 5000** | **5001 - 6000** | **> 6000** |
| --- | --- | --- | --- | --- | --- | --- | --- | --- | --- |
| **Cases** | **26%** | **3%** | **23%** | **17%** | **17%** | **5%** | **3%** | **3%** | **3%** |
| **Deaths** | **25%** | **4%** | **21%** | **21%** | **17%** | **4%** | **4%** | **0%** | **4%** |
| **Discharge** | **28%** | **0%** | **27%** | **9%** | **18%** | **9%** | **0%** | **9%** | **0%** |

## DISCUSSION

This study showed that 58.8% of the admitted patients were men and that of the infections that were diagnosed in the ER and ICU of the Hospital Regional de Gurupi, 76% is equivalent to pneumonia, with 61.8% being older than 60 years with a mortality rate 47.1%. In comparison with a study carried out in Belo Horizonte, more than half of the hospitalizations for pneumonia were in people over 60 years of age, with 64.3% being male^18^. It is observed that this statistic corroborates this research, where the prevalence is found in the male population with community-acquired pneumonia, aged over 60 years.

Although Rivers^19^ mentioned in his article that early antibiotic therapy administered in the first 6 hours of patient admission increases survival, this was not the reality observed in this study, due to the mortality rate found in those first hours of administration and the beginning of antibiotic therapy being 72.7%. As shown in (Table 1), the junction of 0 to 120 ‘and 121 to 360 minutes between hospitalization and the start of drug treatment has a mortality rate of 29%, which means a high number for the number of patients who were hospitalized and started antibiotic therapy at the same time.

Yoshihara explains in a study that mortality in severe sepsis is multifactorial, resulting from the interaction of several factors. The prognosis of sepsis is entirely linked to the fact of both nosocomial and community infection, thus causing dysfunctions in the patient’s system, as gram negative bacteria evolve more severely causing sepsis or septic shock^20^. In this research, it was evidenced that 55.9% of the patients developed sepsis after hospital admission due to community infection, which is mostly a gram-negative bacterium.

The mortality of these patients is extremely alarming due to the number being considered high, exceeding 50% of the patients surveyed. In Primary Care, the diagnosis of infection for clinical purposes is often performed empirically, based on signs and symptoms reported by the patient, and not based on bacterial culture or antibiogram, and when these tests are requested, there is a delay in the result causing possible proliferation. bacterial, in addition to incorrect use of antibiotics making the bacteria resistant to other antimicrobials. In addition, Brazil does not have a community infection surveillance system, which makes it difficult to form a national panorama with the real situation of the problem in question^21^. Thus, it was observed that many of the admitted patients had an exacerbated infectious condition, with serious complications and resistance to antibiotics for treatment, which made it very difficult to treat the patients in question.

## CONCLUSION

This research showed a high mortality rate of included patients who were diagnosed with community infection, mostly composed of elderly people between 70 and 80 years of age. It is worth noting that these arrived at an exacerbated infectious degree of the admitted patients, which makes population management difficult. We also observed that there may be a failure in primary care, which makes an early diagnosis and longer survival of this group of patients difficult. We also analyzed that early antibiotic therapy in patients admitted to ER was not relevant in reducing mortality, since most infections in this group of patients were already in an advanced stage at hospital admission.

## Data Availability

www.datasus.gov.br

